# Relative fertility of HIV-positive women in the ART era: updated estimates from national household survey data

**DOI:** 10.64898/2026.08.11.26360197

**Authors:** Jeffrey W Imai-Eaton, Robert Glaubius, Mary Mahy, Leigh F Johnson, John Stover, Milly Marston

## Abstract

**Objectives:** Estimate fertility rate ratios (FRR) of HIV-positive relative to HIV-negative women in sub-Saharan Africa (SSA) by age group, CD4 stage, ART status, and country.

**Design:** Analysis of nationally representative household surveys with HIV serological testing.

**Methods:** We analysed current pregnancy and births in the past three years by HIV status from 72 nationally-representative household surveys in SSA between 2003 and 2017.

Spectrum 2018 estimates for the distribution by CD4 stage and ART status were used to infer fertility of women on ART from changes in fertility of all HIV-positive women as ART coverage increased. We allowed regional differences in the age pattern of relative fertility and estimated country-specific random effects.

**Results:** The ratio of fertility in untreated HIV-positive women with CD4 ≥500 to HIV-negative women was 1.6 to 1.8 for age 15-19, relatively similar to 10% times lower for age 20-29, and 15–50% lower above age 30. Among age 15-19, each 15-point increase in percent sexually active reduced relative excess fertility by 24%. Fertility decreased with lower untreated CD4 count stages, consistent with previous estimates. Women on ART >6 months had fertility closer to that of HIV-negative women for ages 15-29, but still 25–40% lower above age 30. There was substantial variation across countries.

**Conclusions:** Fertility differences for HIV-positive women compared to HIV-negative women are smaller than previous estimates, but vary substantially across countries. Recent data suggest fertility of women on ART is greater than that of untreated HIV-positive women, but remains lower than HIV-negative women. This conclusion should be reviewed as new evidence becomes available.

## Introduction

Estimates of the HIV prevalence among pregnant women and the number of births to HIV-positive women are central to HIV epidemic estimates in a number of ways, including (1) determining the number of mother-to-child transmissions of HIV and subsequent prevalence of HIV among children and young adults, (2) estimating need for and coverage of prevention of mother-to-child transmission (PMTCT) services, and (3) interpreting trends in HIV prevalence observed among pregnant women attending antenatal care (ANC) for inferring population-wide HIV prevalence and incidence trends.

The Spectrum model uses assumptions about the fertility of HIV-positive women relative to HIV-negative women to generate estimates of the number of pregnancies and births to HIV-positive women. Fertility rate ratios (FRR) allow relative fertility to vary by age, stage of HIV infection, and antiretroviral treatment (ART) status. In sub-Saharan Africa (SSA), it has been consistently documented that among the youngest women, HIV-positive women have greater fertility than HIV-negative women due to selection of sexually active women at risk for both fertility and HIV infection. The relative fertility changes with age and at older ages HIV-positive women have lower fertility than HIV-negative women [1–4]. Multiple factors may contribute to these differences in fertility, including reduced fecundity due to effects of HIV, reduced sexual activity, reduced exposure to pregnancy due to higher levels of widowhood and divorce, and different fertility preferences and contraceptive choices [5].

Each of these determinants of fertility differences for HIV-positive women are likely to change in the context fo ART, and evidence about the fertility of women on ART is sparser. Clinical cohort studies have documented high pregnancy incidence and increased fertility intentions among pregnant women [6,7] and previous modelled estimates have assumed that women on ART have the same fertility as HIV-negative women of the same age. However, a systemic review in 2016 concluded that data were insufficient to characterize the effects of ART on fertility at the population level and identified methodological challenges impeding interpretation of available data [8].

Recent analysis of 49 Demographic and Health Surveys conducted in SSA [4] concluded that (1) overall fertility reductions among HIV-positive women compared to HIV-negative women were not as large as suggested by previous analyses of household survey [3] or population cohort data [9], (2) there are substantial regional and residency differences in relative fertility patterns, and (3) relative fertility of HIV-positive women has increased at higher ART coverage levels, but still remains below that of HIV-negative women, in contrast to model assumptions.

The purpose of this analysis was to further analyze nationally representative household survey data to (1) estimate model fertility rate ratio model parameters consistent with the Spectrum model structure, (2) incorporate recent nationally representative household surveys in SSA, which provide substantial additional evidence about HIV and fertility following the widespread adoption of ‘Option B+’ and earlier ART eligibility for all PLHIV, and (3) combine evidence from outcomes about birth histories [3,4] and current pregnancy by HIV status [10], which have previously been analysed separately.

## Methods

The Spectrum model allows the age-specific fertility rate of untreated HIV-positive women relative to HIV-negative women to vary according to 5-year age group and CD4 count category. For country *c* in year *t*, the fertility rate of untreated HIV-positive women in age group *a* ∈ {15-19, …,45-49}, and CD4 stage of infection *s* ∈ {≥500, 350-499, 250-349, 200-249, 100-199, 50-99, <50} is expressed as

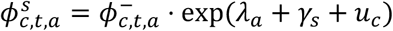

where 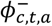 is the fertility rate for HIV-negative women in country *c*, year *t* and age group *a, λ*_*a*_ is the log fertility rate ratio (FRR) for HIV-positive women with CD4 ≥500 compared to HIV-negative women in age group *a, γ*_*s*_ is the log FRR of untreated HIV-positive women in stage *s* relative to women with CD4 ≥500, and *u*_*c*_ is a country-level effect reflecting country-specific variation in the relative fertility of HIV-positive women. For identifiability, *γ*_≥500_ ∶= 1.0.

Among the youngest women aged 15–19 years, the relative fertility of HIV-positive women varies as a function of the country-specific proportion who were sexually active in the past 12 months, as proposed by Chen and Walker [3]. This accounts for strength of selection of sexually active women at risk for both HIV infection and fertility. We parameterised this relationship as

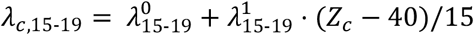

where *Z*_*c*_ is the percentage of 15-19 year-old women reporting sexual activity in the past 12 months, such that 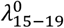 is the log FRR if 40% of 15-19 year-olds are sexually active, approximately the median across SSA countries, and 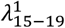 is the change in log FRR per 15 percentage point change in sexually active young women.

For women on ART for fewer than six months, the same relative fertility is assumed as for untreated women since these women would not have been on ART at the time of conception and likely initiated ART during their pregnancy. For women on longer than six months, there is a single parameter 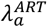 for the log FRR of women on ART compared to HIV-negative women of the same age

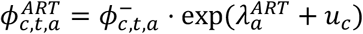

where *u*_*c*_ is the same country-level effect as above for the relative fertility of untreated HIV-positive women.

### Data

We analysed data from 72 nationally-representative household surveys conducted since 2003 in 31 sub-Saharan African countries in which HIV serological status could be linked to individual survey responses about birth histories or current pregnancy status (Figure 1; Table S1). These included 51 Demographic and Health (DHS) surveys, nine AIDS Indicator Surveys (AIS), seven Population HIV Impact Assessment (PHIA) surveys, three Human Sciences Research Council (HSRC) surveys in South Africa, and the Kenya AIDS Indicator Surveys (KAIS) in 2007 and 2012. Sixteen surveys were conducted since 2015. All surveys employed a standard stratified two-stage cluster sampling design and observations were weighted to account for different sampling probabilities and survey participation. We used normalized survey weights that accounted for participation in HIV serological testing as all analyses of fertility by HIV status were restricted to this group of participants.

**Figure 1.**
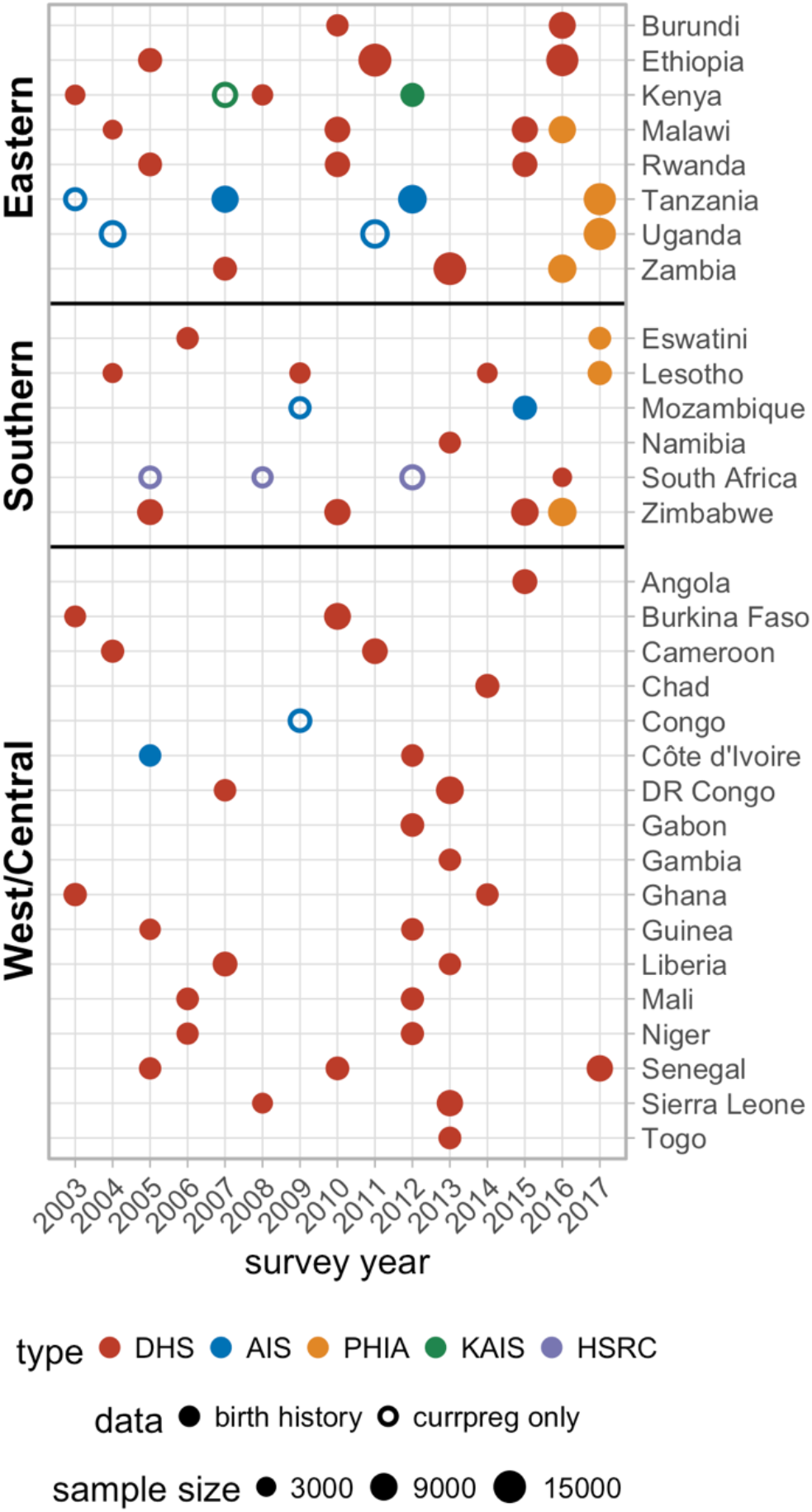
Nationally representative household surveys analysed. Sample size indicates the number of women aged 15-49 who completed individual interview and participated in HIV serological testing.

The modelled outcomes were the fertility rate in the three years before the survey and self-reported pregnancy status at the time of the survey. All 72 surveys included data about self-reported current pregnancy status at the time of the survey. From these surveys, we extracted the total number 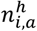 of women survey respondents in survey *i*, by HIV status *h*∈ {HIV-, HIV+}, and age group *a* ∈ {15-19, …,45-49} and the weighted number 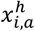 who self-reported being currently pregnant at the time of the survey.

Sixty-three surveys included data about recent births in the years preceding the survey. The 51 DHSs, Cote d’Ivoire 2005 AIS, Mozambique 2015 AIS, and Tanzania 2007 and 2011 AISs recorded full birth histories for all women respondents. From these surveys, we constructed exposure episodes for the three years preceding the survey and calculated the weighted number of person years 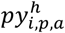 and births 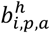 stratified by HIV status *h* at the time of the survey, age-group *a* at the time of exposure and the number of years and time preceding the survey *p* ∈ {1, 2, or 3 years}. Consistent with previous analyses of DHS birth histories to estimate relative fertility by HIV status [3,4], we used the preceding three years to balance maximising the person years of observation included while minimising misclassification of HIV status over the exposure period through measuring HIV status at the time of the survey. The seven PHIA surveys and KAIS 2012 survey only recorded births during the three or two years preceding the survey. For these surveys, we extracted number of person-years and births by HIV status and age group for the one year preceding the survey.

For all 72 surveys, we calculated the percentage of all women respondents aged 15–19 years who reported being sexually active in the past 12 months for use as a covariate predictor of the country-specific FRR *λ*_*c*,15-19_ among this age group, as described above.

### Statistical methods

The purpose of this analysis was to estimate the log FRR parameters 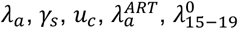 and 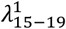 required by Spectrum from population survey data that has informed previous epidemiological analysis of patterns of HIV subfertility by age, calendar time, and region [3,4,10]. The ideal data for such an analysis would be longitudinal data in which it is known when women seroconverted with HIV, progressed through CD4 stages, and initiated ART relative to when they became pregnant. However, from household survey data, the only information available is the women’s HIV status at the time of the survey. In some recent surveys (e.g. PHIA), the CD4 cell count and ARV status are also known at the time of survey, but not at the time of conception of current or earlier pregnancies.

Instead, we estimated the distribution of HIV-positive women by CD4 stage and on ART greater than 6 months in a given country and year using outputs from existing Spectrum 2018 national estimates. We used this distribution to predict the aggregate fertility rate for all HIV-positive women given values for the FRR parameters of interest and fitted this to the observed fertility rates by HIV status from the survey data. Specifically, we extracted the number of HIV-positive women stratified by year, 5-year age group, stage of infection, and ART duration from Spectrum 2018 estimates for all countries in sub-Saharan Africa, then calculated weights 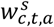 for the proportion of HIV-positive women in age group *a* in year *t* who are untreated or on ART ≤6 months in CD4 stage *s* and 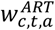 as the proportion of HIV-positive women aged *a* in year *t* who are on ART 6 months or longer, such that

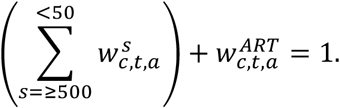

The fertility rate among all HIV-positive women is expressed as

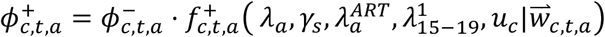

where 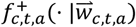 is the weighted average fertility rate ratio for HIV-positive women weighted by the distribution 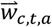 of HIV-positive women across CD4 and treatment stages, given as:

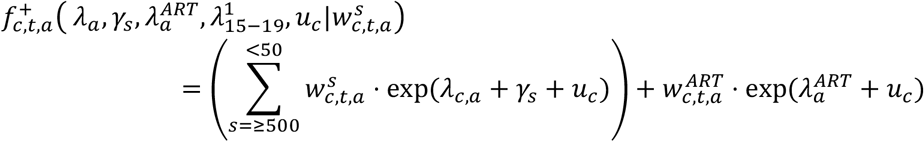

With

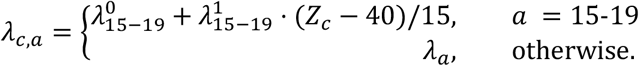

For estimating the model from survey data, we included survey-specific indicator variables 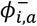 for the ASFR of HIV-negative women. To jointly model both fertility rates from retrospective birth histories and self-reported current pregnancy states, we introduced a variable *β*_*currpre*<_ as the difference in the log probability of reporting being currently pregnant compared to the annual birth rate. If women knew their pregnancy status immediately upon conception, we would expect 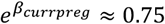 given that pregnancy lasts for nine months, but we expect the estimated value for 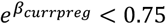 since women may only become aware of and self-report their pregnancy some weeks or months into the pregnancy.

In light of evidence of regional variation in relative fertility of HIV-positive women from Marston *et al*. [4], we introduced regional effects for the age pattern of relative fertility such that *λ*_*a*_ becomes *λ*_*r,a*_ for *r* ∈ {East, Southern, West/Central}. Due to relatively few births to HIV-positive women observed above age 40, we collapsed the age groups to *a* ∈ {15-19, …, 30-34, 35-49}. Finally, also following Marston *et al*., to allow for the fact that HIV status is measured at the time of the survey and the relationship might have been different for births occurring two or three years before the survey, we included an interaction 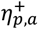 between HIV status, age group, and the number of years *p* ∈ {0,1,2,3} years before the survey, where *p* = 0 was the reference category used for women currently pregnant at the time of the survey, and *p* ∈ {1,2,3} for births history data zero, one, and two years before the survey.

Taken together, we estimated the model:

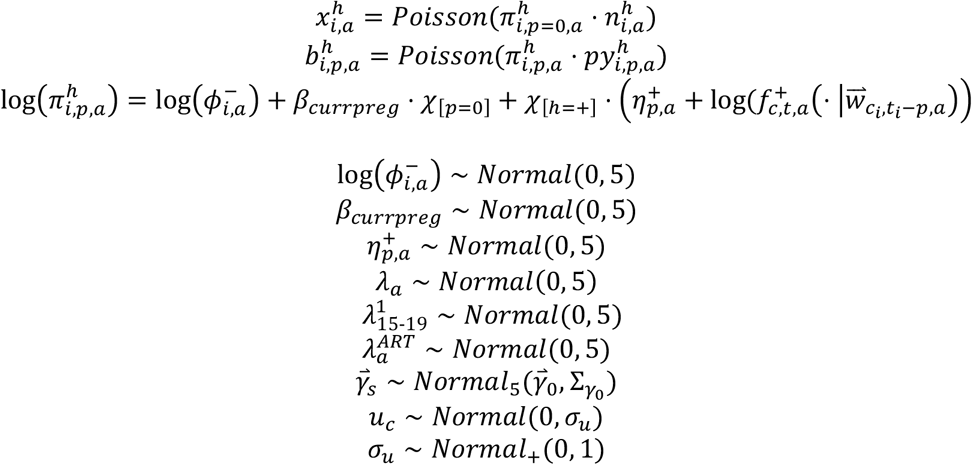

*χ*_[*v*=*x*]_ is an indicator function taking the value 1 if the condition *v* = *x* is met and 0 otherwise to indicate interactions. All regression parameters have diffuse *Normal*(0, 5) prior distributions, except for 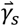, *u*_*c*_ and *σ*_*u*_. Since data about CD4 count at the time of pregnancy or duration of infection are not available, there is little information to inform the FRR by stage of infection *γ*_*s*_ from the cross-sectional survey data. Consequently, we used an informative multivariate normal prior distribution (Table S1) for these parameters based on estimates previously derived for Spectrum from observed patterns of fertility by duration of HIV infection among women in four general population HIV cohort studies in eastern and southern Africa [9]. We use a diffuse standard half-normal distribution for the prior on the standard deviation *σ*_*u*_ of the country-specific effects *u*_*c*_.

### Analysis

DHS and AIS survey data were processed in R using the package *rdhs* and data from other surveys were extracted from relevant survey datasets. Age-specific fertility rates stratified by HIV status were calculated using the package *demogsurv* and the proportion currently pregnant and HIV prevalence were calculated using the *survey* package. We extracted detailed population stratifications by age, sex, CD4 category and treatment duration from Spectrum 2018 estimates by selecting the “Save pop1 array to .xlsx” option in Spectrum and recalculating each projection.

The statistical model was implemented in the statistical computing language *Stan* and we estimated the posterior distribution using No-U-Turn Sampler (NUTS) algorithm as implemented in package *rstan* v2.18.2 [11,12]. We sampled twelve MCMC chains with 1000 iterations each, allocating the first 500 as warm-up and adaptation phase and retaining the second 500, for 6000 total posterior samples.

In addition to fitting the model using all data from all 72 surveys, we conducted several sensitivity analyses. First, we estimated the model using only data from 56 surveys conducted between 2003 and 2014 to understand how FRR estimates are affected by recent surveys since previous analyses [4]. Second, we estimated the model using only birth history survey data [3,4], and only data about current pregnancy status [10].

Finally, we combined posterior estimates for FRR parameters with Spectrum 2018 outputs for the female population, PLHIV, and number of births by age to calculate the expected number of births to HIV-positive women and ASFR by HIV status. We calculated (1) the ratio of the total fertility rate (TFR) among HIV-positive women divided by TFR among HIV-negative women, and (2) the ratio of HIV prevalence among pregnant women versus the HIV prevalence among all women aged 15-49 over the period 1990 to 2020 to understand the implications of the estimated relative fertility patterns on fertility and HIV prevalence among pregnant women over the course of the HIV epidemic.

Computer code for all analyses is available from https://github.com/jeffeaton/spectrum-frr-estimates. The secondary analysis protocol for this study was reviewed and approved by the Imperial College London Research Ethics Committee (ICREC #6461007).

## Results

The 72 nationally representative household surveys reported data about current pregnancy and HIV status for 476,421 women aged 15-49 in 31 SSA countries, including 40,848 HIV-positive women and 435,573 HIV-negative women. Birth history data from 63 surveys consisted of 171,367 births during 1,033,125 person-years. Figure 1 and Supplementary Table S2 describes the data available from each survey and Supplementary Figures S1–S3 illustrate survey estimates for trends in TFR, percentage currently pregnant, and ASFR by HIV status for each country.

### Relative fertility by age, stage of infection, and ART status

Posterior estimates for fertility rate ratio (FRR) parameters are summarised in Table 1 and Figure 2. Among young women aged 15-19, the FRR for untreated HIV-positive women with CD4 >500 (exp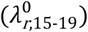) was 1.58 (95% CI 1.25–1.97) in east Africa, 1.69 (1.36–2.09) in southern Africa, and 1.81 (1.42–2.28) in west and central Africa when the proportion sexually active in the past 12 months was the reference value of 40%. Each 15%-point increase in the percent sexually active reduced the FRR by 0.73 (0.66–0.82) times (Figure 2D).

**Table 1.**
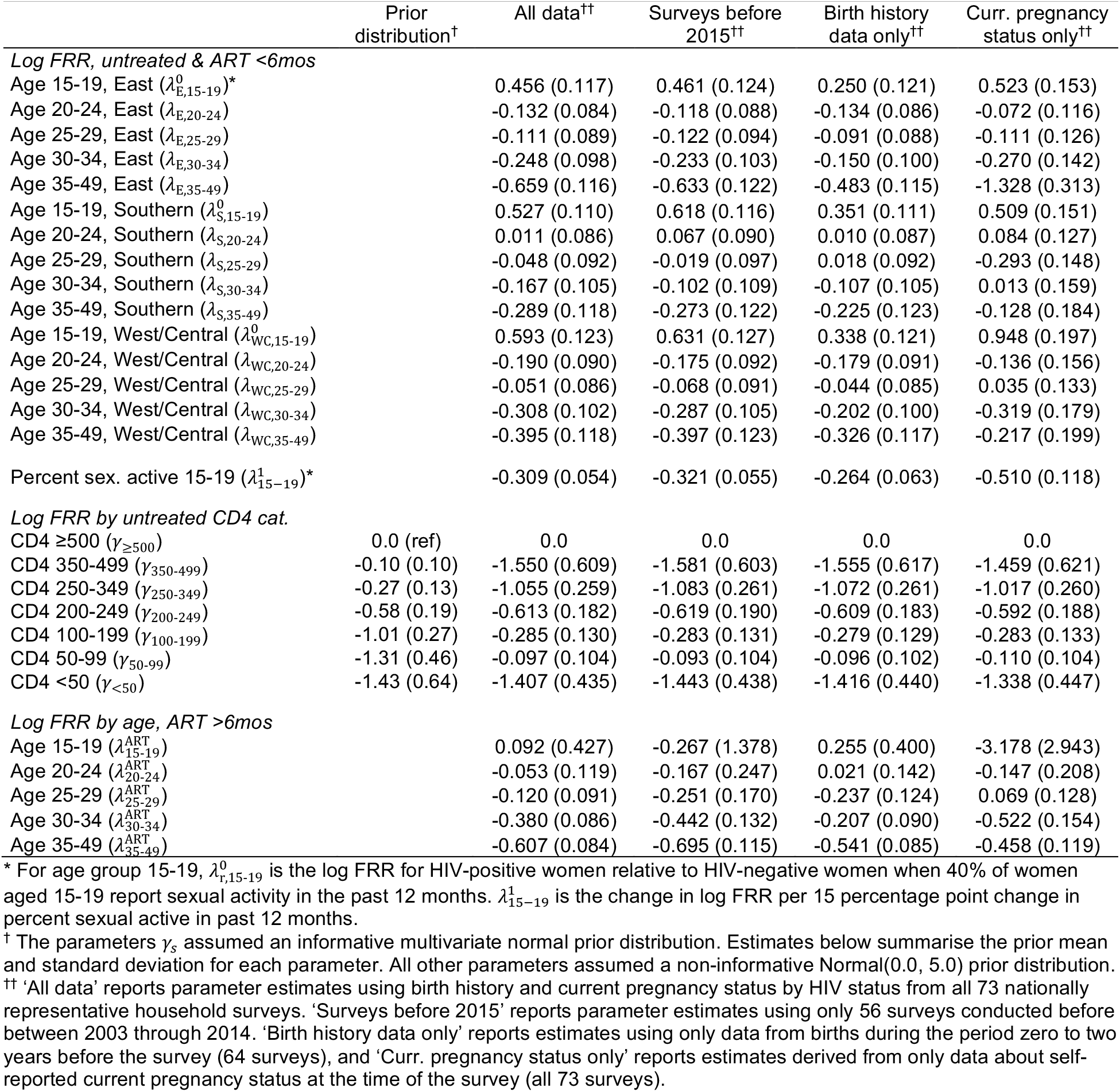
Posterior mean and standard deviation for model parameter estimates.

**Figure 2.**
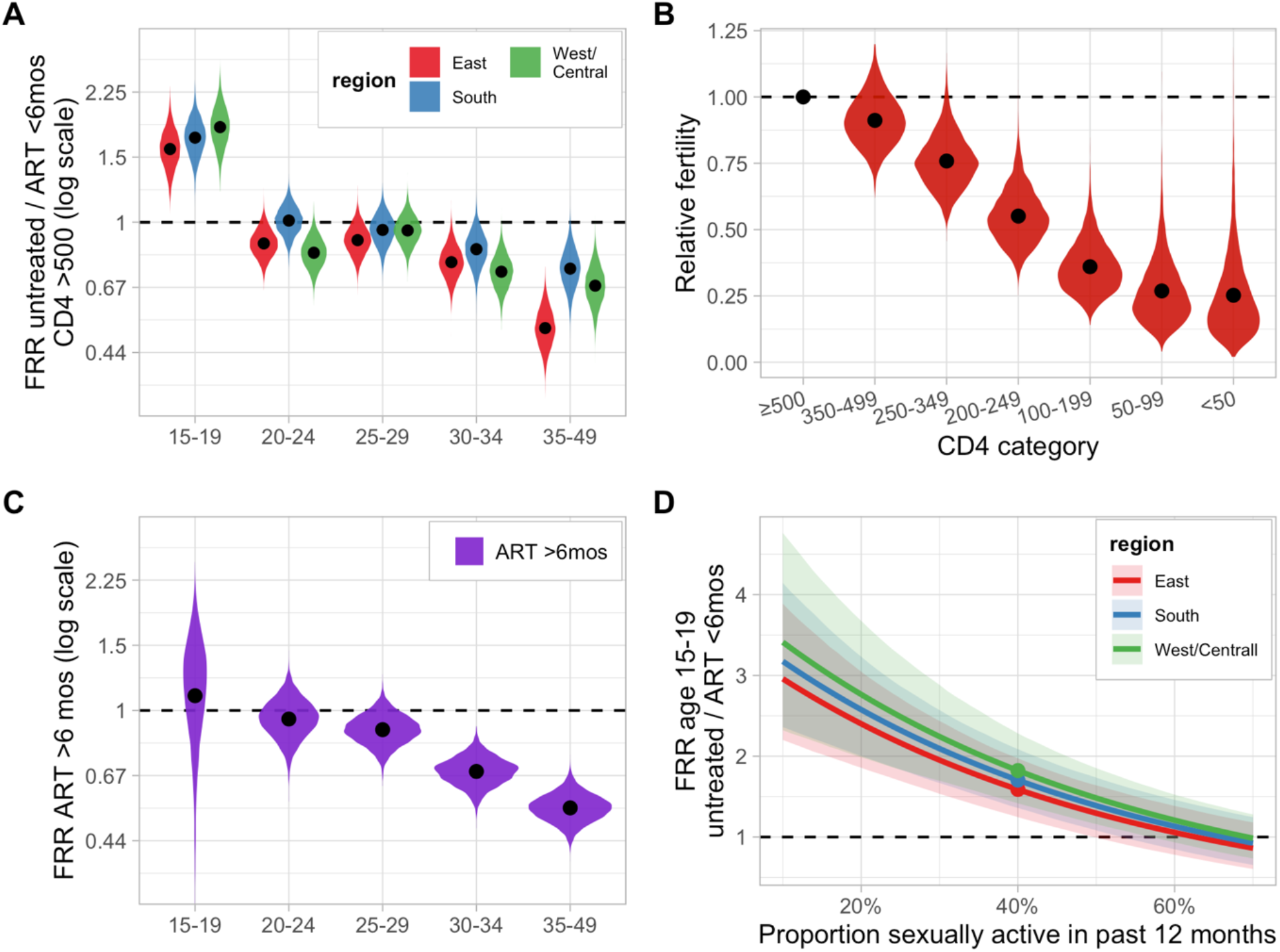
Posterior estimates for FRR parameters. (A) FRR for untreated HIV-positive women with CD4 ≥500 by age relative to HIV-negative women. (B) FRR by CD4 category, relative to untreated HIV-positive women with CD4 ≥500. (C) FRR for women on ART >6 months compared to HIV-negative women. (D) Predicted FRR for age 15-19 age group as a function of the percentage of women aged 15-19 reporting sexual activity in the previous 12 months before the survey. Reference value at 40% corresponds to the FRR for age 15-19 in panel (A). For (A)-(C), points reflect posterior mean estimates and violin plots indicate posterior density estimates. For (D), shaded regions reflects 95% credible intervals.

Among the highest fertility age groups 20-24 and 25-29 years, in southern Africa the fertility of untreated HIV-positive women with CD4 ≥500 was similar to HIV-negative women in southern Africa with FRR of 1.01 (0.85–1.19) and 0.95 (0.79–1.13), respectively. In east Africa and west/central Africa, these women had slightly lower relative fertility at 0.88 (0.74–1.03) and 0.83 (0.69–0.99) for age 20-24 and 0.89 (0.75, 1.06) and 0.95 (0.80–1.12) for age 25-29 years. Above age 30, untreated HIV-positive women with CD4 ≥500 had substantially lower fertility than HIV-negative women, ranging between 15 to 30% lower for age 30-34 and 25 to 50% lower above age 35.

Posterior means for the log FRR by untreated CD4 category *γ*_*s*_ declined steadily with decreasing CD4 stage (Figure 2B). Thus, the overall fertility of untreated HIV-positive women by age will be lower than the age-specific FRR parameters presented in Figure 2A, and this relative relationship will evolve over the course of the HIV epidemic as the CD4 distribution of untreated women evolves with changes in HIV incidence and ART scale up. Posterior mean and standard deviations were similar to the prior means and the standard deviations (Table 1), reflecting that the cross-sectional survey data do not contain much information about relative fertility by duration or stage of HIV infection, as expected.

In contrast to women untreated or on ART <6 months, women aged 15-19 on ART 6 months or longer were estimated to have fertility only 1.10 (0.47–1.99) times than among HIV-negative women of the same age. It makes sense that women on ART aged 15-19 would experience fertility similar to HIV-negative women since a large proportion of them are likely to be long-term survivors of mother-to-child HIV transmission and not necessarily any more likely to be sexually active than HIV-negative women, though this was not mechanistically imposed by the model structure.

Women on ART 6 months or greater aged 20-24 and 25-29 had modestly lower fertility than HIV-negative women with FRRs of 0.95 (0.74–1.19) and 0.89 (0.74–1.06), respectively.

Women above age 30 on ART were still estimated to have substantially lower fertility than HIV-negative women of the same age, with an estimated FRR of 0.68 (0.57–0.81) for age 30-34 and 0.55 (0.46–0.64) for age 35-49.

### Country-level variation

There was substantial variation between countries in the relative fertility of HIV-positive women compared to HIV-negative women (Figure 3). The estimated standard deviation *σ*_*u*_ of the country-specific random effect for log FRR was 0.13 (95% CI 0.08–0.20), implying that the relative fertility of HIV-positive women in any given country is expected to be 10% different from the average effects. Note that these country-specific effects are in addition to the regional variation in relative fertility by age reported in Table 1 and Figure 2A. High HIV prevalence countries with late HIV epidemics in southern Africa such as Eswatini and South Africa had especially high fertility among HIV-positive women and HIV-positive women in Kenya had high fertility relative to other east African countries. HIV-positive women in Ethiopia and Zambia had relatively lower fertility compared to other east African countries.

**Figure 3.**
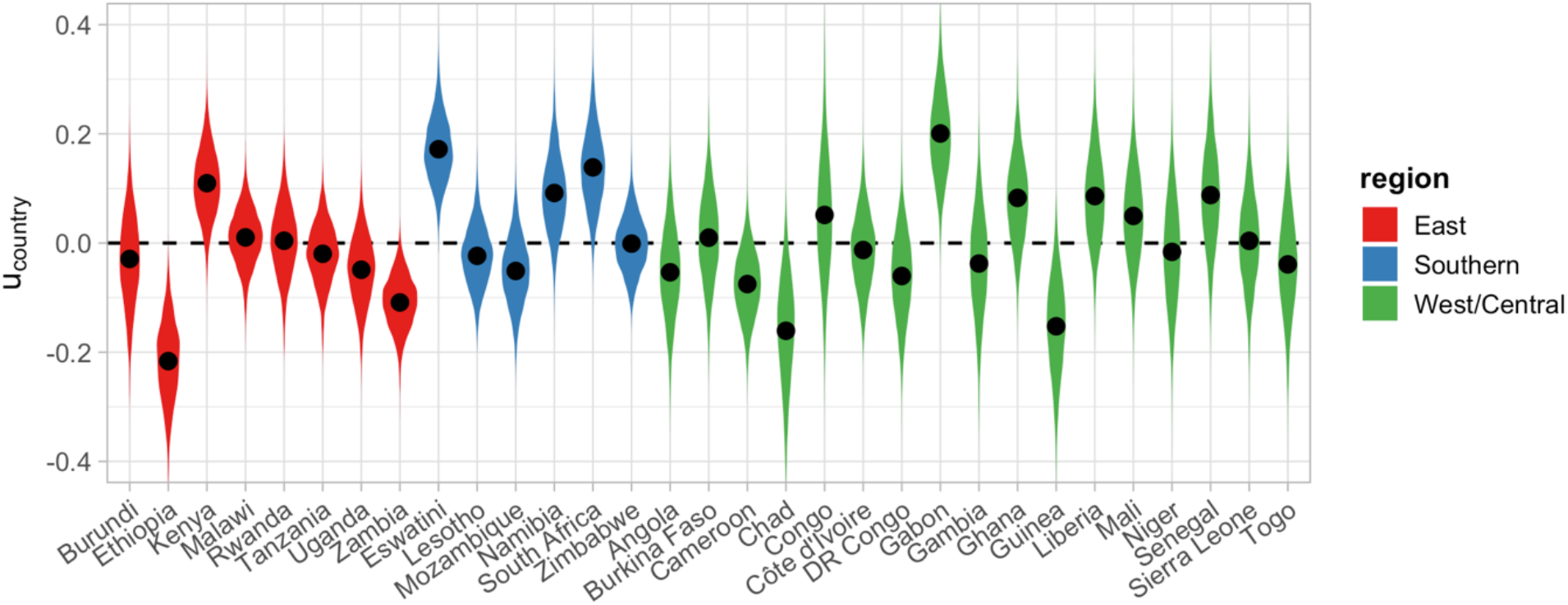
Country-specific random effects for the log relative fertility rate of HIV-positive women compared to HIV-negative women. Random effects are relative to regional average FRR estimates shown in Figure 2A. Points reflect posterior mean and violin plots reflect posterior density.

Uncertainty was large around the country-specific effects, and larger in West/Central African countries with fewer surveys and lower HIV prevalence, resulting in fewer HIV-positive women sampled from which to estimate relative fertility patterns.

### Evidence from recent data

Restricting our analysis to only surveys conducted before 2015 did not substantially affect the point estimates for log FRRs by age or CD4 category (Table 1). The posterior standard deviations were systematically larger, indicating slightly greater uncertainty about estimates given the less data incorporated, but only slightly so.

However, the estimated fertility of women on ART for 6 months or longer were substantially higher and posterior standard deviations were roughly half as large when analysing all surveys compared to only pre-2015 surveys. For example, in the highest fertility ages 20-24, 25-29, and 30-34, point estimates for the fertility of women on ART was 5%, 11% and 32% lower when analysing all data, versus 15%, 22%, and 36% when restricting to pre-2015 surveys.

### Consequences for relative fertility and HIV prevalence over the course of the HIV epidemic

Figure 4A illustrates the ratio of the estimated TFR among HIV-positive divided by the TFR among HIV-negative women over the course of the epidemic. TFR is an age-standardized measure, and so this pattern largely reflects the evolution of relative fertility rates over time. The TFR of HIV-positive women initially declines relative to HIV-negative women as the epidemic matures and a greater proportion of women are in low CD4 stages with low fertility. Then the relative TFR begins increasing again from the late-2000s as the fertility rate among HIV-positive women recovers as ART removes HIV-positive women from the low CD4 stages. Consistent with the FRR estimates (Table 1, Figure 2), HIV-positive women in southern African countries have comparatively higher TFRs and women in east African countries have relatively lower TFRs, but there is wide variation across countries.

**Figure 4.**
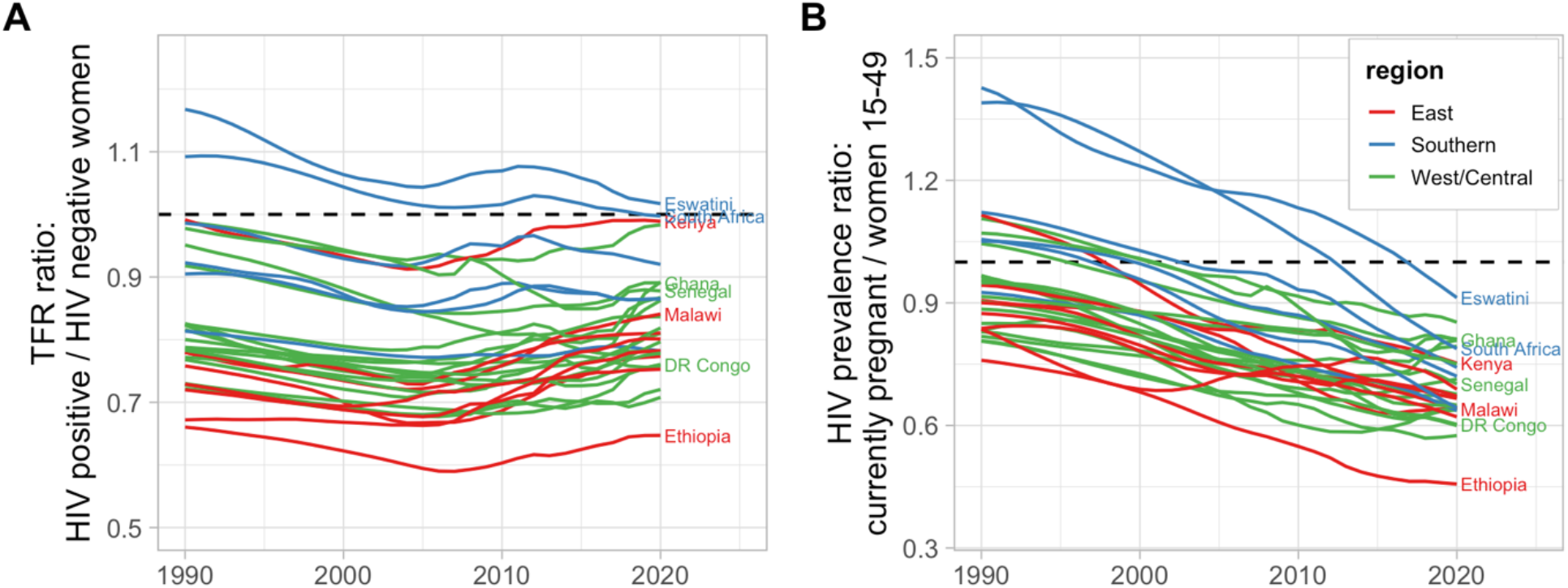
Model predictions for (A) the ratio of TFR among HIV-positive women to HIV-negative women and (B) the ratio of HIV prevalence among currently pregnant women relative to HIV prevalence among all women aged 15–49 years. Estimates for age-specific population size, fertility, and HIV prevalence among all women were taken from Spectrum 2018 estimates and combined with fertility rate ratio estimates shown in Figure 2 and Figure 3. Results reflect posterior mean estimates.

For the HIV prevalence among pregnant women relative to the HIV prevalence of all women aged 15-49 (Figure 4B), the pattern is different. Even while the relative fertility rates of HIV-positive women recovers in the ART era, the ratio of HIV prevalence among pregnant women compared to all women 15-49 continues to decline due to the demographic shift of HIV to older age groups with lower HIV prevalence among younger women resulting from lower HIV incidence and longer survival due to ART leading to increased HIV prevalence at older ages [10].

## Discussion

Our analysis has produced regional estimates for the relative fertility of HIV-positive women by age, CD4 stage, and ART duration in sub-Saharan Africa. These estimates have been used as inputs for revised country HIV estimates in Spectrum 2019. Consistent with recent analysis by Marston *et al*. [4], we found that overall fertility differences for HIV-positive women were somewhat less than previous estimates that have informed Spectrum model assumptions [3,9]. Our model has also for the first-time combined data about birth histories and self-reported current pregnancy by HIV status.

Our analysis estimated that fertility for women on ART >6 months is higher than untreated HIV-positive women, but remains lower than that of HIV-negative women. Sixteen of the 72 surveys analysed were conducted since 2015, reflecting substantial new data that has not been included in previous analyses and covering the era of earlier ART eligibility and ‘Option B+’. Inclusion of these new data substantially increased the precision of the estimates for fertility of women on ART, compared to the data available in previous analyses.

We estimated large cross-country variation in the HIV fertility rate ratio, which should be reflected in country-specific estimates of HIV-positive pregnant women, PMTCT need, and numbers of children infected with HIV. Marston has previously noted the substantial urban/rural differences in both fertility and HIV prevalence in some SSA countries [5], which may confound the relationship between HIV and fertility and explain much of the apparent heterogeneity across countries rather than true differences between countries in the fertility of HIV-positive women. For example, Ethiopia had the largest difference in fertility between HIV-negative and HIV-positive women, and also one of the sharpest urban/rural gradients in both fertility and HIV prevalence. In rural areas, women had TFR of 5.2 and HIV prevalence of only 0.6% but women in urban areas had TFR of 2.3 and HIV prevalence of 3.6% [13]. Further analysis of HIV and fertility at the subnational level and accounting for sociodemographic factors such as education, employment and wealth might further elucidate this relationship.

A major limitation of our analysis is that we did not have directly observed data about pregnancy incidence and fertility by CD4 count and ART status, and instead had to make indirect inference about the fertility of women on ART by correlating temporal changes in the relative fertility of all HIV-positive women with increases in ART coverage. New Population HIV Impact Assessment surveys include survey questions about self-reported date of ART initiation. Further analysis of these data may yield more precision or validation of pregnancy incidence by ART status, though this outcome may be especially sensitive to relatively small reporting errors if they reclassify conceptions as occurring after ART initiation versus initiating ART following HIV diagnosis during pregnancy [14]. Our indirect estimates also relied on outputs from Spectrum 2018 estimates and as such are conditional on the model structure and parameters underpinning those estimates. In particular, if parameters about CD4 distribution or progression are substantially revised, these FRR estimates should also be reevaluated to be consistent.

We encourage caution about extrapolating these estimates due to the ecological nature of our analysis and the rapidly changing context of HIV and fertility in sub-Saharan Africa. They should continue to be reviewed as new data become available. Analysis of differences in sexual behavior and contraceptive use by HIV status by Marston *et al*. has suggested that the lower fertility of HIV-positive women at older ages may be much more attributable to behavioral differences rather than biological effects of HIV infection [5], and consequently this relationship could further evolve as partnership dissolution and widowhood further reduce with mature ART programmes. Mechanistically capturing these dynamics might improve projections about future changes in fertility of HIV-positive women.

Lastly, the uncertainty ranges around our estimates for relative fertility parameters were large and we observed substantial heterogeneity across countries. This suggests that the modelling approach of applying fertility rate ratios to overall population fertility and HIV estimates will likely yield insufficiently precise estimates for numbers of HIV-positive pregnant women to meet current policy objectives—for example reliably distinguishing between 85% to 95% coverage of PMTCT services necessary for Elimination of mother to child transmission validation. Recognizing this, Spectrum has also added a tool to calibrate the country-specific random effect such that the prevalence among pregnant women matches observed prevalence from routine testing of pregnant women at antenatal care [CITE Stover *et al*, this supplement]. This should yield more specific and precise estimates in a given setting. Future work should continue to develop direct and indirect approaches to estimating PMTCT coverage through survey and routine programmatic data.

## Data Availability

Secondary data analysed in this study are available upon online request from the Demographic and Health Survey (DHS) programme, Population HIV Impact Assessment (PHIA) surveys, South Africa Human Sciences Research Council (HSRC), Kenya national data archive.

https://dhsprogram.com/data/available-datasets.cfm

https://phia-data.icap.columbia.edu/datasets

https://hsrc-repository.figshare.com/

https://statistics.knbs.or.ke/nada/index.php

## Acknowledgements

JWE-E, RG, MM, LFJ, JS, and MM conceived the study. All authors contributed to the design of the study. JWI-E and MM analysed survey data. JWI-E, RG, and JS conducted modelling analyses. All authors contributed to drafting and revision of the manuscript for critical content.

This research was supported by UNAIDS, the National Institute of Allergy and Infectious Diseases (NIAID) of the National Institutes of Health under award number R03-AI125001, and the MRC Centre for Global Infectious Disease Analysis (reference MR/X020258/1), funded by the UK Medical Research Council (MRC). This UK funded award is carried out in the frame of the Global Health EDCTP3 Joint Undertaking. The findings and conclusions in this report are those of the author(s) and do not necessarily represent the official position of the funding agencies.

We thank Sasi Jonnalagadda, Peter Young, Paul Stupp, Andrew Voetsch for input on this analysis and guidance on interpretation of PHIA household survey data and participants of the UNAIDS Reference Group on Estimates, Modelling, and Projections for input on previous versions of this manuscript.

We dedicate this manuscript to Prof Basia Zaba, who tirelessly guided us to study the demographic impacts of HIV with passion, vigour, and tenacity.

---

**Figure S1.**
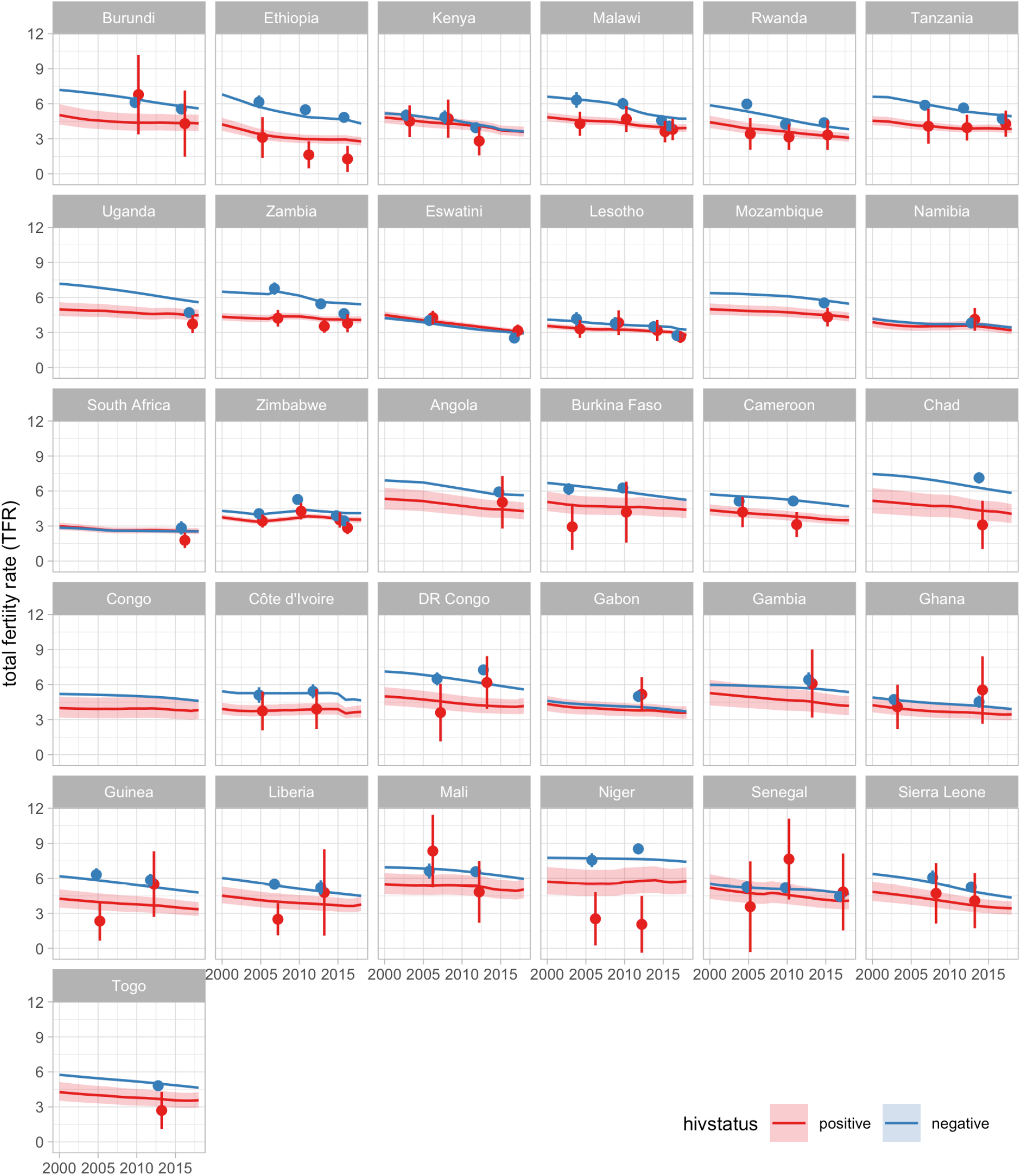
Predicted and observed total fertility rate (TFR) by HIV status. Points and vertical lines reflect direct survey estimates and 95% confidence intervals for TFR by HIV status in the 12 months preceding the survey from 72 household surveys analysed (Table 1). Estimates for total TFR (both HIV-postive and HIV-negative) and age-specific HIV prevalence are taken from Spectrum 2018 point estimates; and estimates for relative fertility of HIV positive women are from the present analysis.

**Figure S2.**
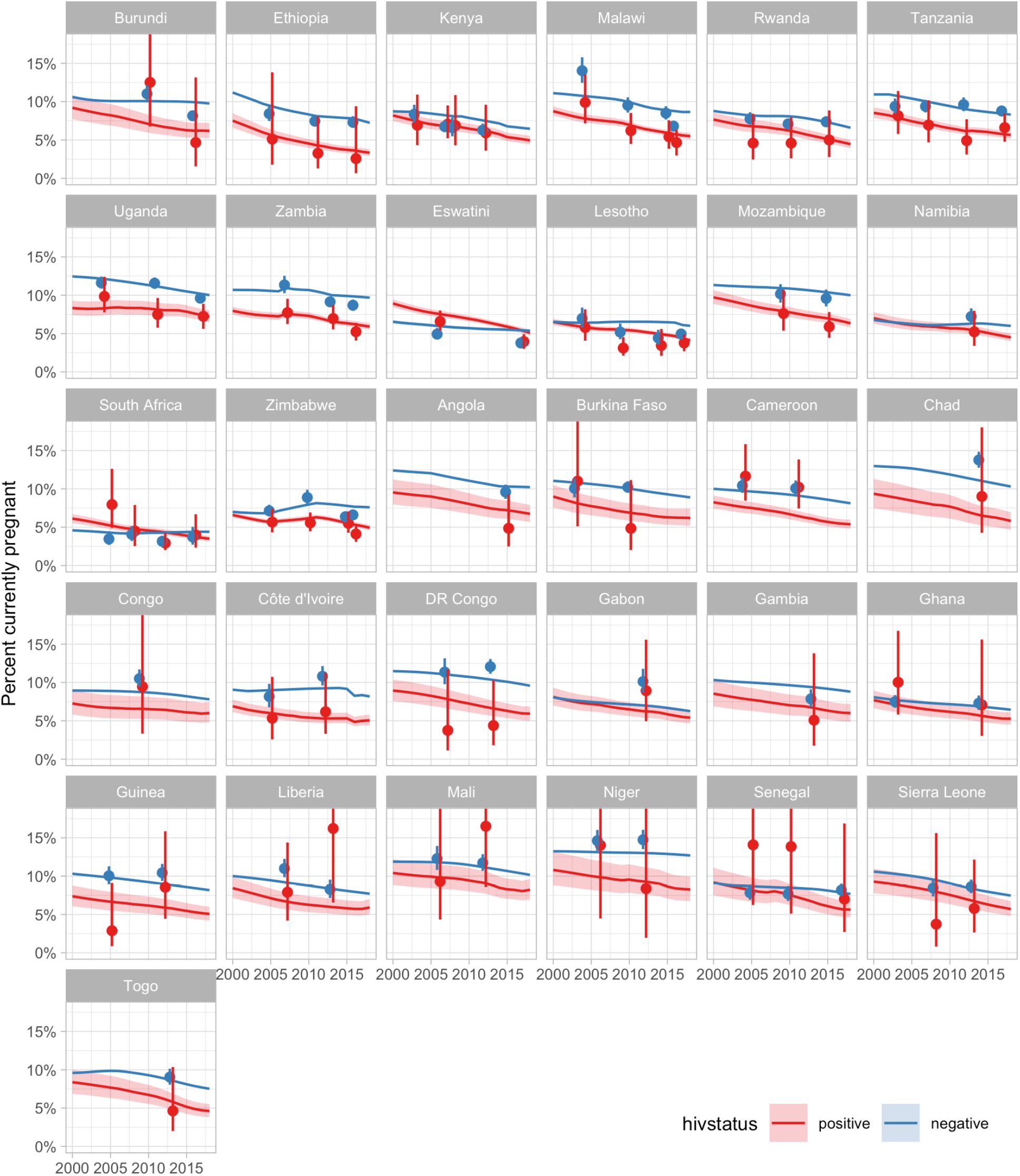
Predicted and observed percent currently pregnant by HIV status. Points and vertical lines reflect direct survey estimates and 95% confidence intervals for the percentage of women self-reporting being currently pregnant at the time of survey by HIV status from 72 household surveys analysed (Table 1). Estimates for total ASFR (both HIV-postive and HIV-negative) and age-specific HIV prevalence are taken from Spectrum 2018 estimates. Estimates for relative fertility of HIV positive women are from the present analysis.

**Figure S3.**
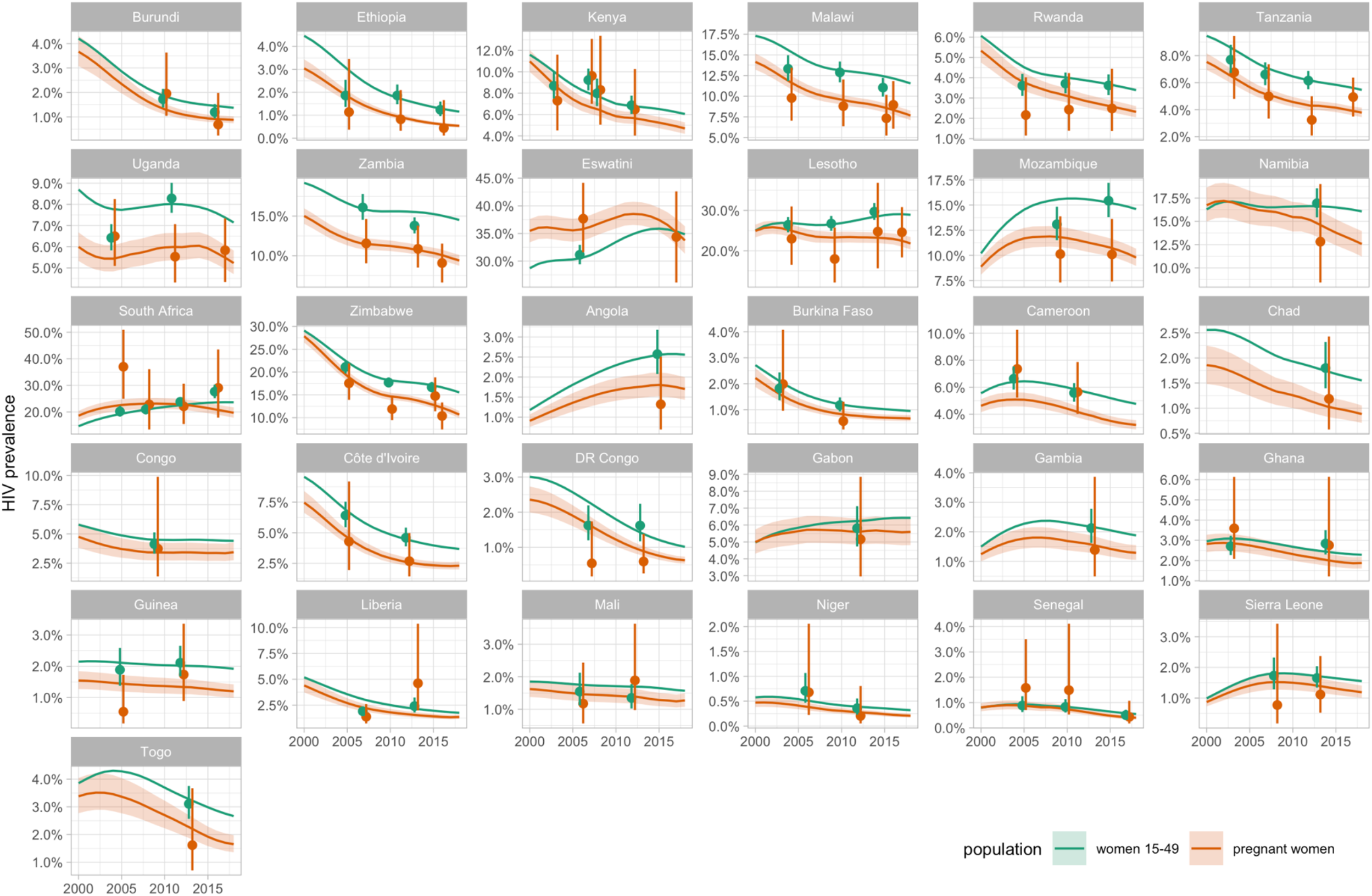
Predicted and observed HIV prevalence among all women aged 15-49 years and currently pregnant women. Points and vertical lines reflect direct survey estimates and 95% confidence intervals for HIV prevalence among all women aged 15-49 and women self-reporting being currently pregnant. Estimates for prevalence age 15-49 are taken from Spectrum 2018 estimates; estimates for prevalence among pregnant women are calculated from relative fertility rates estimated in the present analysis.

